# A Bayesian Re-analysis of the Steroids to Reduce Systemic Inflammation after Infant Heart Surgery (STRESS) Trial

**DOI:** 10.1101/2025.02.10.25322035

**Authors:** Kevin D. Hill, Jake Koerner, Hwanhee Hong, Jennifer S. Li, Christoph Hornik, Prince J. Kannankeril, Jeffrey P. Jacobs, H. Scott Baldwin, Marshall L. Jacobs, Eric M. Graham, Brian Blasiole, David F. Vener, Adil S. Husain, S. Ram Kumar, Alexis Benscoter, Eric Wald, Tara Karamlou, Andrew H Van Bergen, David Overman, Pirooz Eghtesady, Ryan Butts, John S. Kim, John P. Scott, Brett R. Anderson, Michael F. Swartz, Sean M. O’Brien

## Abstract

**Background:** Prophylactic steroids are often used to reduce the systemic inflammatory response to cardiopulmonary bypass in infants undergoing heart surgery. The STRESS trial found that the likelihood of a worse outcome did not differ between infants randomized to methylprednisolone (n=599) versus placebo (n=601) in a risk-adjusted primary analysis (adjusted odds ratio [OR], 0.86; 95% CI, 0.71 to 1.05; P=0.14). However, secondary analyses showed possible benefits with methylprednisolone. To ensure that a potentially efficacious therapy is not unnecessarily avoided, we re-analyzed the STRESS trial using Bayesian analytics to assess the probability of benefit.

**Methods:** Our Bayesian analysis used the original STRESS trial primary outcome measure, a hierarchically ranked composite of death, transplant, major complications and post-operative length of stay. We evaluated probability of benefit (OR<1) versus harm (OR>1) by comparing the posterior distribution of the OR assuming a neutral probability of benefit versus harm with weak prior belief strength (nearly non-informative prior distribution). Reference results were calculated under the vague prior distribution. To convey magnitude of effect we used model parameters to calculate a predicted risk of death, transplant or major complications for methylprednisolone and placebo. Analyses consisted of 10 Markov Chain Monte Carlo simulations, each consisting of 2000 iterations with a 1000 iteration burn-in to ensure proper posterior convergence. Sensitivity analyses evaluated pessimistic (5%-30% prior likelihood of benefit), neutral and optimistic (70%-95%) prior beliefs, and controlled strength of prior belief as weak (30% variance), moderate (15%) and strong (5%).

**Results:** In primary analysis, the posterior probability of benefit from methylprednisolone was 91% and probability of harm was 9%. Composite death or major complication occurred in 18.8% of trial subjects with an absolute risk difference of -2% (95% CI -3%, +1%) associated with methylprednisolone. Each of 9 sensitivity analyses demonstrated greater probability of benefit than harm in the methylprednisolone group with 8 of 9 demonstrating >80% probability of benefit and ≥1% absolute difference in risk of death, transplant or major complications.

**Conclusion:** Probability of benefit with prophylactic methylprednisolone is high and harm is unlikely. This more in-depth analysis of the data expands the initial clinical evaluation of methylprednisolone provided by the STRESS trial.

## Background

Perioperative glucocorticoids are commonly administered to infants undergoing cardiac surgery to mitigate the systemic inflammation caused by cardiopulmonary bypass. This post-bypass systemic inflammatory response contributes to post-operative morbidity and mortality. However, early pediatric trials and adult trials evaluating safety and efficacy of perioperative glucocorticoids have yielded conflicting results^1–10^. The STeroids to REduce Systemic Inflammation after infant heart Surgery (STRESS) Trial randomized 1200 infants (under 1 year of age) undergoing heart surgery with cardiopulmonary bypass to receive either perioperative methylprednisolone or placebo^11,12^. In a covariate-adjusted analysis of the primary endpoint (a ranked composite), the likelihood of a worse outcome did not differ significantly between the methylprednisolone and placebo groups (adjusted odds ratio, 0.86; 95% confidence interval [CI], 0.71 to 1.05; P=0.14)^13^. However, secondary analyses, including an unadjusted analysis of the primary outcome and a win ratio analysis, favored the methylprednisolone group, suggesting potential benefits^13^.

Bayesian analysis offers a unique perspective in evaluating clinical trial data. Unlike frequentist statistical designs that rely on a pre-specified p-value cut-off to determine trial outcomes, Bayesian analyses evaluate the probability of benefit^14^. This approach allows for incorporating prior knowledge to assess the probability of benefit under different hypotheses and with varying degrees of confidence in the prior knowledge^14^. Bayesian analysis is particularly valuable in rare diseases, including most pediatric conditions, where underpowering is common, and where underpowered studies could be discarded despite offering important information. For example, investigators used Bayesian analysis to re-evaluate the Therapeutic Hypothermia after In-Hospital Cardiac Arrest in Children (THAPCA) Trial. Although the original THAPCA analysis did not meet its primary endpoint^15^, Bayesian re-analysis demonstrated a high probability that the intervention offered benefit with a very low likelihood of harm^16^.

We sought to apply Bayesian analysis, including incorporating a range of potential “prior beliefs”, to re-assess the outcomes of the STRESS trial. Our primary objective was to determine the probability of benefit versus harm associated with prophylactic methylprednisolone in order to better guide clinical decision-making.

## Methods

The STRESS Trial was a multi-center double-blind, randomized, controlled “trial within a registry”. Study design and results have been previously reported^11,13^. Briefly, 1263 infants (under 1 year of age) undergoing heart surgery with cardiopulmonary bypass at 24 centers in the United States were randomized to methylprednisolone (30mg/kg administered into the CPB pump prime) versus placebo. Trial outcomes were collected from the Society of Thoracic Surgeons Congenital Heart Surgery Database. The primary outcome was a hierarchically ranked composite endpoint with components ranked commensurate with their perceived clinical significance. Individual components included operative mortality, heart transplant, or any of 13 individual major complications. Outcomes in the composite endpoint were ranked into 97 levels of clinical prioritization as follows: operative death was ranked as 97 (worst outcome); heart transplantation during hospitalization as 96; permanent dialysis, tracheostomy, or neurologic deficit at discharge as 95; postoperative mechanical circulatory support or unplanned cardiac reoperation (exclusive of reoperation for bleeding) as 94; reoperation for bleeding, unplanned delayed sternal closure, or postoperative unplanned interventional cardiac catheterization as 93; postoperative cardiac arrest, multisystem organ failure, kidney failure with temporary dialysis, or postoperative mechanical ventilator support for more than 7 days as 92; and postoperative length of hospital stay of 91 days or longer as 91. Patients with none of these postoperative complications were assigned ranked outcomes according to postoperative length of hospital stay (1 to 90 days). The distribution of the ranked outcome components of the primary endpoint were compared between the trial groups with the use of a proportional-odds logistic-regression model for ordered categorical data with prespecified covariates adjustment for age, weight, prematurity status, and the 2020 STS–European Association for Cardio-Thoracic Surgery (STAT) Mortality Category^17^; in addition, adjustment was made for the random effect of enrollment site.

### Bayesian statistical framework

Bayesian analysis is a statistical framework for determining the relative likelihood of different possible numerical estimates of unknown parameters in light of study data and prior information. It integrates prior beliefs or knowledge about the parameters of interest, encoded as prior distributions, with evidence from the observed data as quantified by the likelihood function. This integration provides a posterior distribution of the parameters of interest, reflecting updated beliefs about the treatment effects, accounting for both the prior information and the evidence from the study. Bayesian analysis provides not only point estimates but also probabilities-based assessment metrics, offering direct numerical quantifications of uncertainty. This makes it particularly well-suited for comparative effectiveness studies, where summarizing evidence and making decisions under uncertainty is critical.

### Statistical analysis

A detailed description of the statistical approach is provided in the Supplementary Methods. Briefly, we performed a Bayesian re-analysis of the STRESS trial’s primary outcome using statistical methods aligned with the original main results presentation^13^. To match the trial’s original analysis, we used data from the same cohort of randomized subjects who received study drug (methylprednisolone or placebo) and were included in the modified intention-to-treat population (n = 599 methylprednisolone, n = 601 placebo). To further align with the original analysis, we used a proportional odds (also known as cumulative logits) and adjusted for the same set of pre-specified covariates, including age, weight, prematurity status, the 2020 STS– European Association for Cardio-Thoracic Surgery (STAT) Mortality Category^17^, and randomizing site. The proportional odds model is an extension of binary logistic regression to accommodate ordered outcomes with more than two categories. The model’s dependent (outcome) variable was an ordered categorical variable representing the patient’s ranked outcome category (1=best, 97=worst). The treatment effect of methylprednisolone versus placebo was expressed as an odds ratio with an odds ratio <1 favoring methylprednisolone and an odds ratio >1 favoring placebo. Additionally, to help interpret the odds ratio’s magnitude, we transformed parameter estimates from the fitted proportional odds model to the scale of probabilities and used this transformation to calculate the difference in the model-predicted probability of death, transplant or major complications for methylprednisolone minus placebo, denoted as “risk difference”. This transformed treatment effect parameter was not estimated in the original results presentation but was included here to make analysis results more clinically interpretable^18^.

### Parameter Estimation

Bayesian analysis requires assigning prior distributions to model parameters reflecting prior knowledge or belief about them. Because our prior information was limited, our goal was to select a prior distribution that would allow inferences to be driven by the study data as opposed to strong prior beliefs. In keeping with this goal, we specified that the logarithm of the odds ratio would follow a normal distribution with mean = 0 and SD = 3. Other model parameters besides the odds ratio were assigned non-informative prior distributions. To assess whether results were sensitive to the choice of prior, we performed sensitivity analyses re-fitting the model using a range of priors chosen to reflect different possible beliefs about the odds ratio’s direction and magnitude, as suggested by Wijeysundera et al. and Zampieri et al^19,20^. Moreover, following Harhay et. al.^16^, we considered priors based on combinations of three possible “best guesses” for the unknown odds ratio, ranging from optimistic (best guess = 0.80; suggesting benefit from methylprednisolone) to pessimistic (best guess = 1.25; suggesting harm from methylprednisolone), and three choices for the strength of belief in this best guess, ranging from weak to strong prior belief (see Table 1 and Supplementary Methods for details). Parameter estimation was performed using Markov Chain Monte Carlo (MCMC) sampling as implemented in the R software package brms^21^. Posterior summaries are based on 10,000 MCMC samples drawn after a burn-in of 10,000 iterations. Convergence of the MCMC procedure was confirmed through evaluation of trace plots, autocorrelation plots, and effective sample size^22^.

**Table 1.**
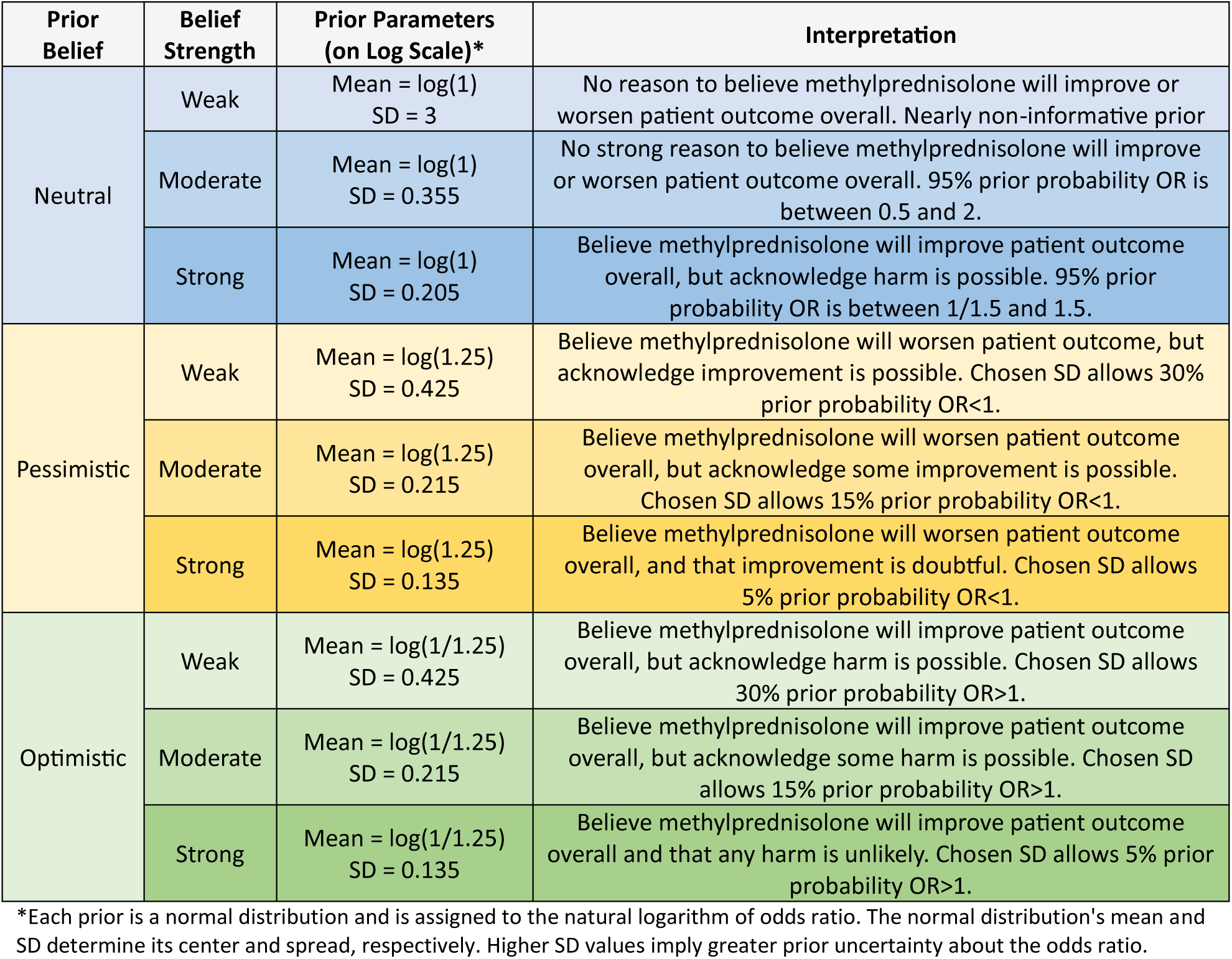
Priors assigned to the odds ratio (OR) in Bayesian re-analysis of STRESS trial. Priors and text interpretations are adapted from Harhay et al^16^. and Zampieri et al^20^. The primary outcome was based on neutral prior belief with weak belief strength.

| Prior Belief | Belief Strength | Prior Parameters (on Log Scale)* | Interpretation |
| --- | --- | --- | --- |
| Neutral | Weak | Mean = $\log(1)$<br>SD = 3 | No reason to believe methylprednisolone will improve or worsen patient outcome overall. Nearly non-informative prior |
| | Moderate | Mean = $\log(1)$<br>SD = 0.355 | No strong reason to believe methylprednisolone will improve or worsen patient outcome overall. 95% prior probability OR is between 0.5 and 2. |
| | Strong | Mean = $\log(1)$<br>SD = 0.205 | Believe methylprednisolone will improve patient outcome overall, but acknowledge harm is possible. 95% prior probability OR is between 1/1.5 and 1.5. |
| Pessimistic | Weak | Mean = $\log(1.25)$<br>SD = 0.425 | Believe methylprednisolone will worsen patient outcome, but acknowledge improvement is possible. Chosen SD allows 30% prior probability OR<1. |
| | Moderate | Mean = $\log(1.25)$<br>SD = 0.215 | Believe methylprednisolone will worsen patient outcome overall, but acknowledge some improvement is possible. Chosen SD allows 15% prior probability OR<1. |
| | Strong | Mean = $\log(1.25)$<br>SD = 0.135 | Believe methylprednisolone will worsen patient outcome overall, and that improvement is doubtful. Chosen SD allows 5% prior probability OR<1. |
| Optimistic | Weak | Mean = $\log(1/1.25)$<br>SD = 0.425 | Believe methylprednisolone will improve patient outcome overall, but acknowledge harm is possible. Chosen SD allows 30% prior probability OR>1. |
| | Moderate | Mean = $\log(1/1.25)$<br>SD = 0.215 | Believe methylprednisolone will improve patient outcome overall, but acknowledge some harm is possible. Chosen SD allows 15% prior probability OR>1. |
| | Strong | Mean = $\log(1/1.25)$<br>SD = 0.135 | Believe methylprednisolone will improve patient outcome overall and that any harm is unlikely. Chosen SD allows 5% prior probability OR>1. |

\*Each prior is a normal distribution and is assigned to the natural logarithm of odds ratio. The normal distribution's mean and SD determine its center and spread, respectively. Higher SD values imply greater prior uncertainty about the odds ratio.

## Results

For the primary outcome, as in the original trial, 1200 trial participants were evaluated (599 patients in the methylprednisolone group and 601 in the placebo group). The cumulative number of endpoint events was lower in the methylprednisolone group than in the placebo group for all successive endpoint categories (Table 2).

**Table 2.** Cumulative endpoint event rates.

| Primary cumulative endpoint groupings | Methylprednisolone<br>N=599 | Placebo<br>N=601 |
| --- | --- | --- |
| Rank = 97 | 12/599 (2.0%) | 17/601 (2.8%) |
| Rank ≥ 96 | 15/599 (2.5%) | 24/601 (4.0%) |
| Rank ≥ 95 | 19/599 (3.2%) | 32/601 (5.3%) |
| Rank ≥ 94 | 63/599 (10.5%) | 68/601 (11.3%) |
| Rank ≥ 93 | 75/599 (12.5%) | 98/601 (16.3%) |
| Rank ≥ 92 | 99/599 (16.5%) | 120/601 (20.0%) |
| Rank ≥ 91 | 103/599 (17.2%) | 122/601 (20.3%) |

97 - Operative Mortality; 96 - Heart Transplant; 95 - Renal failure with permanent dialysis, neurologic deficit persistent at discharge, or respiratory failure requiring tracheostomy; 94 - Post-operative mechanical circulatory support or unplanned cardiac reoperation (exclusive of reoperation for bleeding); 93 - Reoperation for bleeding, unplanned delayed sternal closure, or post-op unplanned interventional cardiac catheterization; 92 - Post-op cardiac arrest, multi-system organ failure, renal failure with temporary dialysis, or prolonged ventilator support (> 7 days); 91 - Post operative length of stay > 90 days; 1-90 – post-op length of stay in days

In primary risk-adjusted Bayesian analysis (neutral prior belief, weak belief strength), the posterior probability of any benefit (OR<1.0) from methylprednisolone was 91% and the probability of any harm (OR>1.0) was 9%. The methylprednisolone group had a mean odds ratio of 0.87 denoting a 13% mean reduction in the odds of a worse outcome (Table 3, Figure 1). The composite of death or major complication occurred in 17.2% of subjects randomized to methylprednisolone and 20.3% of those randomized to placebo (adjusted odds ratio 0.83; 95% CI, 0.61 to 1.13). In primary risk-adjusted Bayesian analysis, the methylprednisolone group demonstrated a 2% mean reduction in the risk of death, transplant or major complication (Table 4, Figure 1).

**Figure 1:**
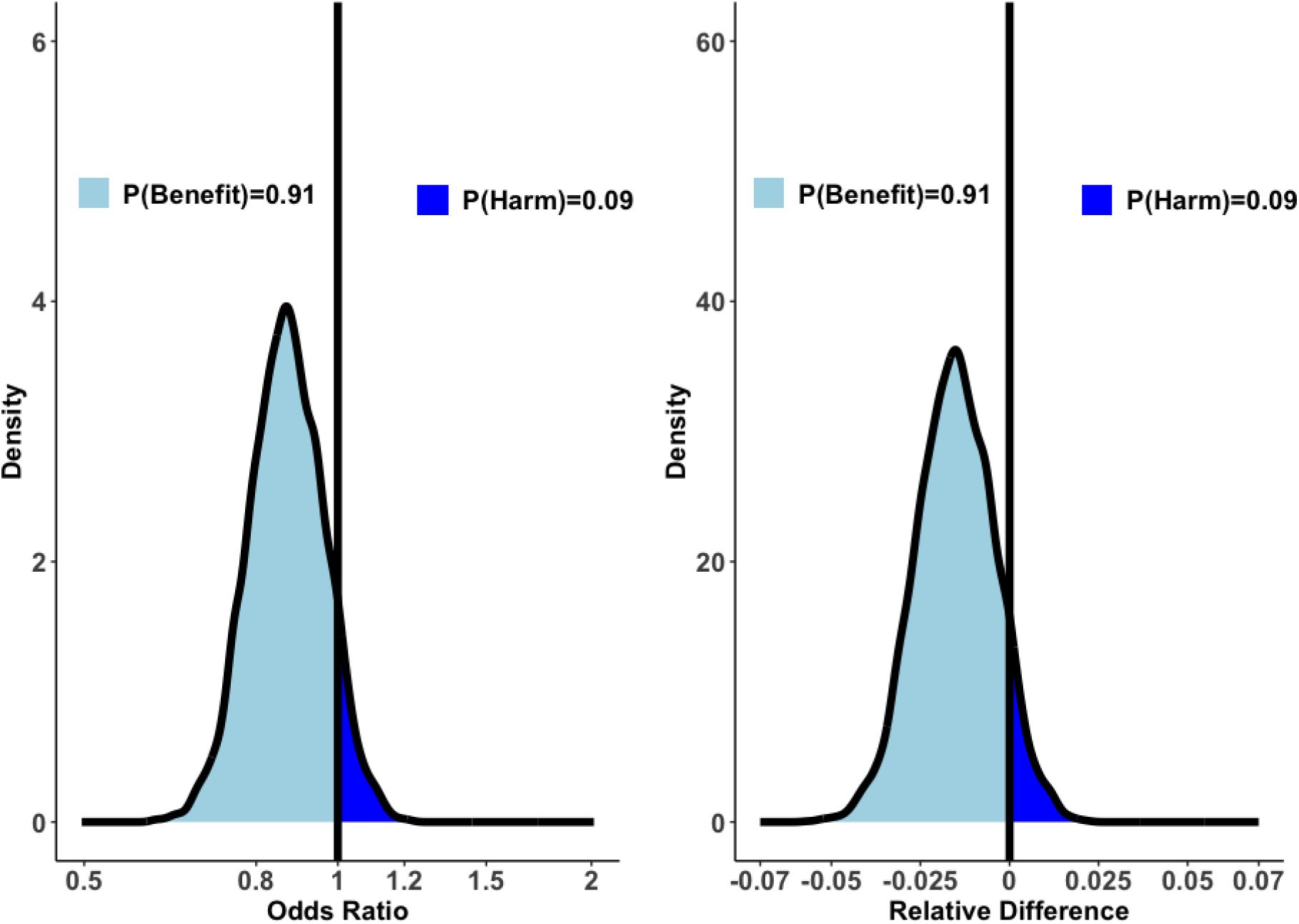
Posterior distributions of odds ratio (OR, left) and risk difference (RD, right) of death, heart transplant or major complications (outcome of rank 90 or higher) under the Neutral, Weak prior setting as defined in Table 1, based on Bayesian analysis of STRESS trial. Light blue indicates region of posterior density for which we expect beneficial patient outcome as the result of being given methylprednisolone (OR<1 or RD<0), whereas dark blue indicates region of posterior for which harm is expected (OR>1 or RD>0).

**Table 3:** Posterior summary of odds ratio (OR) with 95% credible intervals given in parenthesis, as well as posterior probability of OR being less than 0.8, 0.9, 1, and greater than 1, 1.1, 1.25, under different prior settings as defined in. **Table 1, based on Bayesian analysis of STRESS trial.**

| Prior Belief | Belief Strength | Estimate (95% CI) | Prob of OR<0.8 | Prob of OR<0.9 | Prob of OR<1 | Prob of OR>1 | Prob of OR>1.1 | Prob of OR>1.2 |
| --- | --- | --- | --- | --- | --- | --- | --- | --- |
| Neutral | Weak | 0.87 (0.71, 1.06) | 0.22 | 0.65 | 0.92 | 0.08 | 0.01 | <0.01 |
|  | Moderate | 0.88 (0.72, 1.06) | 0.18 | 0.62 | 0.91 | 0.09 | 0.01 | <0.01 |
|  | Strong | 0.89 (0.74, 1.07) | 0.13 | 0.55 | 0.90 | 0.10 | 0.01 | <0.01 |
| Pessimistic | Weak | 0.89 (0.73, 1.07) | 0.16 | 0.57 | 0.89 | 0.11 | 0.01 | <0.01 |
|  | Moderate | 0.93 (0.77, 1.11) | 0.06 | 0.38 | 0.80 | 0.20 | 0.03 | <0.01 |
|  | Strong | 0.99 (0.84, 1.16) | <0.01 | 0.12 | 0.57 | 0.43 | 0.09 | 0.01 |
| Optimistic | Weak | 0.87 (0.71, 1.04) | 0.22 | 0.67 | 0.93 | 0.07 | 0.01 | <0.01 |
|  | Moderate | 0.86 (0.71, 1.02) | 0.25 | 0.72 | 0.96 | 0.04 | <0.01 | <0.01 |
|  | Strong | 0.84 (0.72, 0.99) | 0.27 | 0.80 | 0.98 | 0.02 | <0.01 | <0.01 |

**Table 4:**
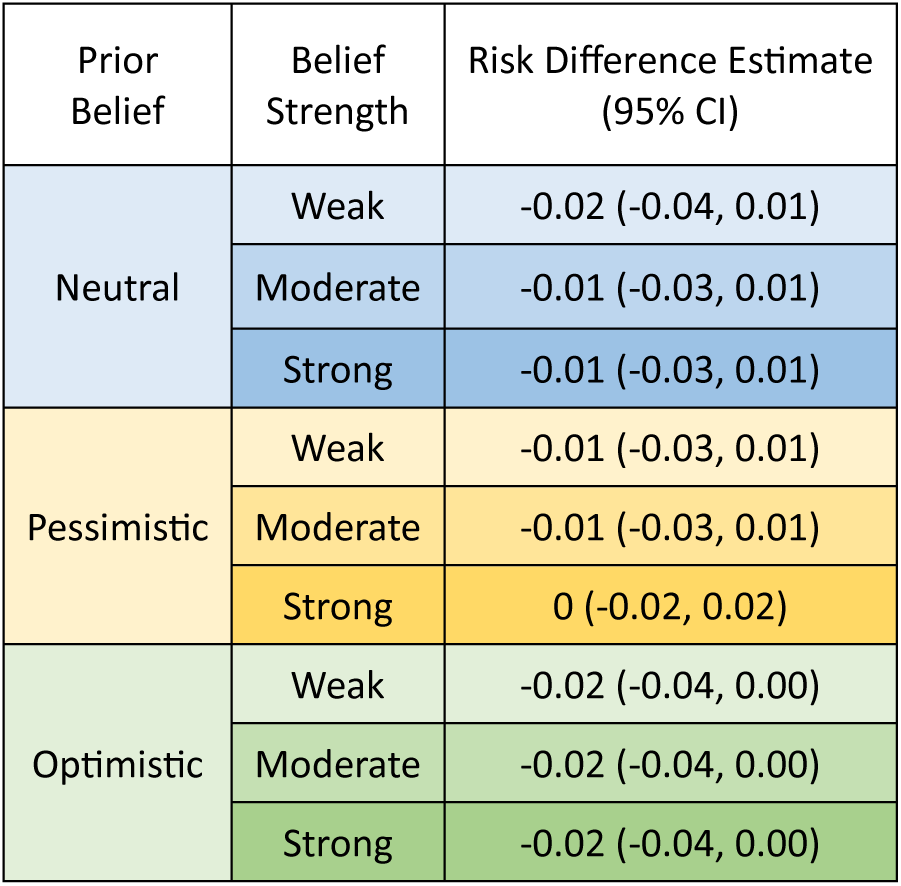
Posterior summary of risk difference (RD) of death, heart transplant or major complications (outcome in category 91 or higher) with 95% credible intervals given in parenthesis, as well as posterior probability of RD being less than -0.05, -0.025, 0, and greater than 0, 0.025, 0.05, under different prior settings as defined in. **Table 1, based on Bayesian analysis of STRESS trail.**

| Prior Belief | Belief Strength | Risk Difference Estimate (95% CI) |
| --- | --- | --- |
| Neutral | Weak | -0.02 (-0.04, 0.01) |
|  | Moderate | -0.01 (-0.03, 0.01) |
|  | Strong | -0.01 (-0.03, 0.01) |
| Pessimistic | Weak | -0.01 (-0.03, 0.01) |
|  | Moderate | -0.01 (-0.03, 0.01) |
|  | Strong | 0 (-0.02, 0.02) |
| Optimistic | Weak | -0.02 (-0.04, 0.00) |
|  | Moderate | -0.02 (-0.04, 0.00) |
|  | Strong | -0.02 (-0.04, 0.00) |

In sensitivity analyses, evaluating a range of prior beliefs and belief strengths (Table 3,4, Figure 2), the posterior probability of benefit favored methylprednisolone in all scenarios. The mean reduction in the odds of a worse outcome ranged from 1% (pessimistic prior belief with strong belief strength) to 15% (optimistic prior belief with strong belief strength). The absolute risk reduction for death or major complications was ≥1% for 8/9 scenarios and there was no scenario where the average risk of death or major complications was increased in the methylprednisolone group.

**Figure 2:**
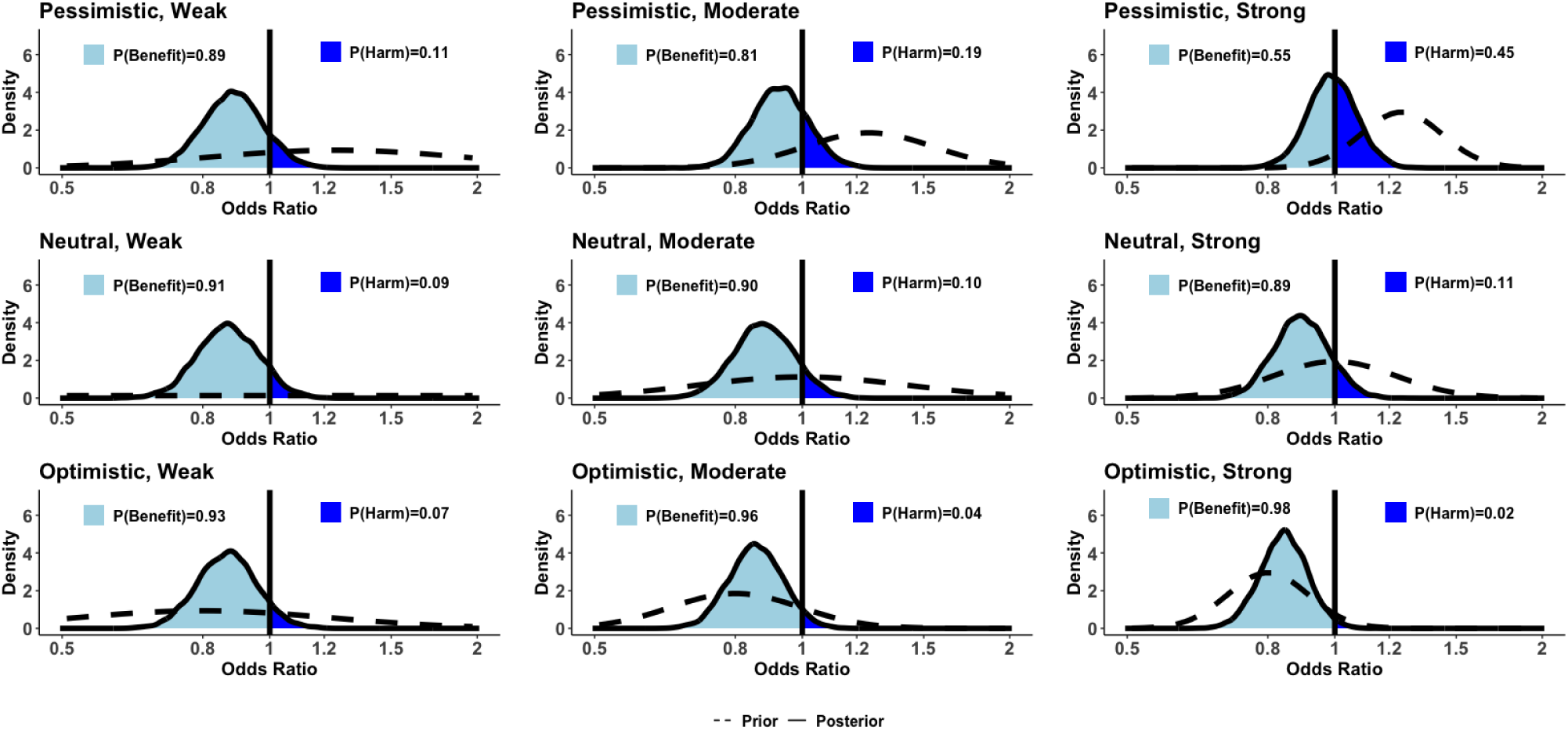
Density plots of the odds ratio (OR) under different prior settings as defined in Table 1, based on Bayesian analysis of STRESS trial. Light blue indicates region of posterior density for which we expect beneficial patient outcome as the result of being given methylprednisolone (OR<1), whereas dark blue indicates region of posterior for which harm is expected (OR>1).

## Discussion

Our secondary Bayesian re-analysis of the STRESS trial demonstrates high probability that perioperative glucocorticoids offer some benefit to infants undergoing cardiopulmonary bypass with a low likelihood of harm. Our results were consistent across a range of prior beliefs and belief strengths, showing a 9-14% overall reduction in the odds of a worse outcome and a 1-2% absolute reduction in the risk of death or major complications for all but the most pessimistic and most optimistic belief scenarios.

Previous trials of perioperative glucocorticoids in children have yielded inconsistent results. The STRESS Trial, the largest glucocorticoid trial to date, found no statistically significant difference between the glucocorticoid and placebo arms (adjusted odds ratio, 0.86; 95% confidence interval [CI], 0.71 to 1.05; P=0.14)^13^. However, at each level of the hierarchically ranked endpoint, results favored the methylprednisolone arm and the confidence interval skewed in favor of methylprednisolone. Two pre-specified secondary analyses, one without covariate adjustment and the other using a win ratio, also favored the methylprednisolone arm. Similar results were observed in two other relatively large contemporary glucocorticoid trials. The DECISION trial^4^, which randomized 394 infants from Brazil, China, and Russia to dexamethasone versus placebo, found no significant difference in the primary composite outcome. However, all individual components of the composite favored the dexamethasone group, with the 95% confidence intervals skewed in favor of dexamethasone (absolute risk reduction, 7.4%; 95% CI, −0.8% to 15.3%; P = .20). Graham et al. randomized 190 neonates to methylprednisolone versus placebo^2^ and found that the primary composite outcome occurred less frequently in the methylprednisolone group (33% versus 42%), with all individual components of the primary outcome less frequent in the methylprednisolone group and the 95% confidence interval skewed towards methylprednisolone (odds ratio [OR]: 0.63; 95% CI: 0.31 to 1.30; p = 0.21). However, the study findings did not reach statistical significance. A meta-analysis of 12 glucocorticoid trials performed between 2000 and 2023, including all three of the aforementioned trials (n = 2132 subjects), found no mortality benefit but did show that glucocorticoids decreased the duration of mechanical ventilation and the incidence of post-operative low cardiac output syndrome^8^.

Taken together, these trials and meta-analyses consistently favor glucocorticoids, but none have reached statistical significance for the primary endpoints. Traditionally, three “negative” trials and a negative meta-analysis would be considered strong evidence against glucocorticoid efficacy. However, the “gold standard” p-value threshold can lead to the inadvertent discarding of therapies that may offer clinical benefit because the frequentist analytic design favors the null hypothesis unless there is near-irrefutable evidence (p<0.05) that the null hypothesis is wrong^23^. The p value obtained from a frequentist analysis is intentionally conservative, reflecting the probability of observing a treatment effect if the null hypothesis is correct and there is no treatment effect. In contrast, a Bayesian calculation assesses the probability of a pre-specified treatment effect, incorporating prior beliefs or evidence into the analytic framework.^24^ This approach mirrors clinical thinking, where new evidence is weighed in the context of existing evidence to arrive at the best possible therapeutic decision. For these reasons and because investigators recognize the limitations of a frequentist approach in some trials^25^ Bayesian approaches are increasingly used to analyze randomized controlled trial data^16,26,27^. Technological advances have further increased the feasibility of running the complex simulations required for these analyses.

Bayesian analyses are particularly suited to trials where the adverse consequences of an erroneous conclusion are small. In the case of perioperative steroids, a frequentist interpretation sets a high threshold (p<0.05) for rejecting the null hypothesis (i.e., that there is no treatment benefit) and thereby minimizes the chances of “doing harm” by rejecting the null hypothesis. However, the risk-benefit assessment should consider past experience with the drug, including both the possibility of efficacy and the potential for drug-related adverse events. Perioperative steroids have a well-established safety profile due to decades of experience and multiple prior trials. Transient hyperglycemia is the most common side effect seen with higher doses and has been demonstrated in both pediatric and adult trials^1,2,4,6,13^. Other side effects are rare and have not impacted outcomes. Because of this well-established safety profile, it is arguably more useful for providers to understand both the true probability of benefit and the magnitude of benefit to make a well-informed clinical decision on the drug’s overarching clinical utility. Our data suggest that perioperative glucocorticoids offer a high probability of efficacy, and a clinically meaningful estimated reduction in the absolute likelihood of death, heart transplant or major complications of approximately 2%. Notably, in the STRESS trial, death, heart transplant or major complications occurred in 17.2% of subjects randomized to methylprednisolone and 20.3% of those randomized to placebo, a risk difference of 3.1%. Thus, our Bayesian analysis provides a conservative estimate of the risk difference, perhaps reflecting a difference in baseline risk factors between the placebo and methylprednisolone subjects. Irrespective, our Bayesian analysis indicates that continued use of prophylactic methylprednisolone may be justified despite prior “negative” trials. Notably, prior data indicate that lower glucocorticoid doses may be equally efficacious as the 30 mg/kg methylprednisolone dose administered in the STRESS trial^28^. Potentially, lower doses may minimize adverse effects (such as transient hyperglycemia) without impacting the efficacy profile.

### Limitations

Our Bayesian assessment of harm associated with glucocorticoids focuses on the potential for harm with respect to the components of the primary STRESS trial composite outcome measure. Harm can also be measured in terms of drug side effects that do not impact outcomes. We did not re-analyze the potential for harmful side effects, including hyperglycemia and infection.

Hyperglycemia is a well-established side effect of glucocorticoids demonstrated in the STRESS trial and multiple other RCTs^1,2,4,6,13^. Our intent was not to discount this side effect but rather to provide insight into the probability of benefit with respect to the outcomes that providers hope to impact when prescribing glucocorticoids. These benefits must be considered in the context of the known side effect profile of glucocorticoids. Notably, although hyperglycemia did occur more frequently in the methylprednisolone group, our results show that hyperglycemia did not contribute to worse outcomes, indicating that if there is an adverse impact of hyperglycemia on outcomes, it is perhaps mitigated by the beneficial effects of methylprednisolone. The potential for increased infection risk with glucocorticoids is less clear. Prior registry analyses have indicated a possible increased risk^29^ but RCTs and meta-analyses have not shown any signal^2,4,8–10,13^. We did not apply Bayesian analytics to evaluate the probability of infection associated with glucocorticoids in the STRESS trial because event rates were very similar between the two trial arms (n=43/599, 7.2% versus n=38/601, 6.3% with infectious complications for methylprednisolone versus placebo, respectively), and because we did not have sufficiently strong prior evidence to help guide our analytic approach.

In conclusion, Bayesian analysis can provide unique insights into the probability of benefit and can augment frequentist interpretations of a clinical trial. Our Bayesian re-analysis of the STRESS trial indicates a high probability that perioperative methylprednisolone can offer some clinical benefit.

## Data Availability

A STRESS Trial deidentified dataset will be available one year after publication

## References

1. Dieleman JM, Nierich AP, Rosseel PM, et al. Intraoperative high-dose dexamethasone for cardiac surgery: a randomized controlled trial. JAMA 2012;308(17):1761–7. DOI: 10.1001/jama.2012.14144.

2. Graham EM, Martin RH, Buckley JR, et al. Corticosteroid Therapy in Neonates Undergoing Cardiopulmonary Bypass: Randomized Controlled Trial. J Am Coll Cardiol 2019;74(5):659–668. DOI: 10.1016/j.jacc.2019.05.060.

3. Li Y, Luo Q, Wu X, Jia Y, Yan F. Perioperative Corticosteroid Therapy in Children Undergoing Cardiac Surgery: A Systematic Review and Meta-Analysis. Front Pediatr 2020;8:350. DOI: 10.3389/fped.2020.00350.

4. Lomivorotov V, Kornilov I, Boboshko V, et al. Effect of Intraoperative Dexamethasone on Major Complications and Mortality Among Infants Undergoing Cardiac Surgery: The DECISION Randomized Clinical Trial. JAMA 2020;323(24):2485–2492. DOI: 10.1001/jama.2020.8133.

5. Scrascia G, Rotunno C, Guida P, et al. Perioperative steroids administration in pediatric cardiac surgery: a meta-analysis of randomized controlled trials*. Pediatric critical care medicine: a journal of the Society of Critical Care Medicine and the World Federation of Pediatric Intensive and Critical Care Societies 2014;15(5):435–42. DOI: 10.1097/PCC.0000000000000128.

6. Whitlock RP, Devereaux PJ, Teoh KH, et al. Methylprednisolone in patients undergoing cardiopulmonary bypass (SIRS): a randomised, double-blind, placebo-controlled trial. Lancet 2015;386(10000):1243–1253. DOI: 10.1016/S0140-6736(15)00273-1.

7. Whitlock RP, Dieleman JM, Belley-Cote E, et al. The Effect of Steroids in Patients Undergoing Cardiopulmonary Bypass: An Individual Patient Meta-Analysis of Two Randomized Trials. J Cardiothorac Vasc Anesth 2020;34(1):99–105. DOI: 10.1053/j.jvca.2019.06.012.

8. Takeshita J, Nakajima Y, Tachibana K, Takeuchi M, Shime N. Efficacy of perioperative prophylactic administration of corticosteroids in pediatric cardiac surgeries using cardiopulmonary bypass: a systematic review with meta-analysis. Anaesth Crit Care Pain Med 2023;42(6):101281. DOI: 10.1016/j.accpm.2023.101281.

9. Chen D, Du Y. Analysis of perioperative corticosteroid therapy in children undergoing cardiac surgery: A systematic review and meta-analysis. Clin Cardiol 2023;46(6):607–614. DOI: 10.1002/clc.24018.

10. Bronicki RA, Flores S, Loomba RS, et al. Impact of Corticosteroids on Cardiopulmonary Bypass Induced Inflammation in Children: A Meta-Analysis. Ann Thorac Surg 2021;112(4):1363–1370. DOI: 10.1016/j.athoracsur.2020.09.062.

11. Hill KD, Baldwin HS, Bichel DP, et al. Rationale and design of the STeroids to REduce Systemic inflammation after infant heart Surgery (STRESS) trial. Am Heart J 2020;220:192–202. DOI: 10.1016/j.ahj.2019.11.016.

12. Hill KD, Baldwin HS, Bichel DP, et al. Overcoming underpowering: Trial simulations and a global rank end point to optimize clinical trials in children with heart disease. Am Heart J 2020;226:188–197. DOI: 10.1016/j.ahj.2020.05.011.

13. Hill KD, Kannankeril PJ, Jacobs JP, et al. Methylprednisolone for Heart Surgery in Infants - A Randomized, Controlled Trial. N Engl J Med 2022;387(23):2138–2149. DOI: 10.1056/NEJMoa2212667.

14. Berry DA. Bayesian clinical trials. Nat Rev Drug Discov 2006;5(1):27–36. DOI: 10.1038/nrd1927.

15. Moler FW, Silverstein FS, Holubkov R, et al. Therapeutic Hypothermia after In-Hospital Cardiac Arrest in Children. N Engl J Med 2017;376(4):318–329. DOI: 10.1056/NEJMoa1610493.

16. Harhay MO, Blette BS, Granholm A, et al. A Bayesian Interpretation of a Pediatric Cardiac Arrest Trial (THAPCA-OH). NEJM Evid 2023;2(1):EVIDoa2200196. DOI: 10.1056/EVIDoa2200196.

17. Jacobs ML, Jacobs JP, Thibault D, et al. Updating an Empirically Based Tool for Analyzing Congenital Heart Surgery Mortality. World J Pediatr Congenit Heart Surg 2021;12(2):246–281. DOI: 10.1177/2150135121991528.

18. King G TM, Wittenberg J. Making the most of statistical analyses: Improving interpretation and presentation. American journal of political science 2000;44(2):341–55.

19. Wijeysundera DN, Austin PC, Hux JE, Beattie WS, Laupacis A. Bayesian statistical inference enhances the interpretation of contemporary randomized controlled trials. J Clin Epidemiol 2009;62(1):13–21 e5. DOI: 10.1016/j.jclinepi.2008.07.006.

20. Zampieri FG, Casey JD, Shankar-Hari M, Harrell FE, Jr., Harhay MO. Using Bayesian Methods to Augment the Interpretation of Critical Care Trials. An Overview of Theory and Example Reanalysis of the Alveolar Recruitment for Acute Respiratory Distress Syndrome Trial. Am J Respir Crit Care Med 2021;203(5):543–552. DOI: 10.1164/rccm.202006-2381CP.

21. P B. brms: An R Package for Bayesian Multilevel Models Using Stan. Journal of Statistical Software 2017;80(1):1–28.

22. Gelman A CJ, Stern HS, Rubin DB. Bayesian data analysis. Chapman and Hall/CRC, 1995.

23. Altman DG, Bland JM. Absence of evidence is not evidence of absence. BMJ 1995;311(7003):485. DOI: 10.1136/bmj.311.7003.485.

24. Ferguson J. Bayesian interpretation of p values in clinical trials. BMJ Evid Based Med 2022;27(5):313–316. DOI: 10.1136/bmjebm-2020-111603.

25. Amrhein V, Greenland S, McShane B. Scientists rise up against statistical significance. Nature 2019;567(7748):305–307. DOI: 10.1038/d41586-019-00857-9.

26. Lammers D, Richman J, Holcomb JB, Jansen JO. Use of Bayesian Statistics to Reanalyze Data From the Pragmatic Randomized Optimal Platelet and Plasma Ratios Trial. JAMA Netw Open 2023;6(2):e230421. DOI: 10.1001/jamanetworkopen.2023.0421.

27. Maron DJ, Hochman JS, Reynolds HR, et al. Initial Invasive or Conservative Strategy for Stable Coronary Disease. N Engl J Med 2020;382(15):1395–1407. DOI: 10.1056/NEJMoa1915922.

28. Hornik CP, Gonzalez D, Dumond J, et al. Population Pharmacokinetic/Pharmacodynamic Modeling of Methylprednisolone in Neonates Undergoing Cardiopulmonary Bypass. CPT Pharmacometrics Syst Pharmacol 2019;8(12):913–922. DOI: 10.1002/psp4.12470.

29. Pasquali SK, Hall M, Li JS, et al. Corticosteroids and outcome in children undergoing congenital heart surgery: analysis of the Pediatric Health Information Systems database. Circulation 2010;122(21):2123–30. DOI: 10.1161/CIRCULATIONAHA.110.948737.

